# A participatory modelling approach for investigating the spread of COVID-19 in countries of the Eastern Mediterranean Region to support public health decision-making

**DOI:** 10.1101/2021.02.10.21251474

**Authors:** Keyrellous Adib, Penelope A. Hancock, Aysel Rahimli, Bridget Mugisa, Fayez Abdulrazeq, Noha Farag, Ricardo Aguas, Lisa White, Rana Hajjeh, Lubna Al Ariqi, Pierre Nabeth

## Abstract

Early on in the COVID-19 pandemic, the WHO Eastern Mediterranean Regional Office (WHO EMRO) recognised the importance of epidemiological modelling to forecast the progression of the COVID-19 pandemic to support decisions guiding the implementation of response measures. We established a modelling support team to facilitate the application of epidemiological modelling analyses in the Eastern Mediterranean Region (EMR) countries. Here we present an innovative, stepwise approach to participatory modelling of the COVID-19 pandemic that engaged decision-makers and public health professionals from countries throughout all stages of the modelling process. Our approach consisted of first identifying the relevant policy questions, collecting country-specific data, and interpreting model findings from a decision-maker’s perspective, as well as communicating model uncertainty. We used a simple modelling methodology that was adaptable to the shortage of epidemiological data, and the limited modelling capacity, in our region. We discuss the benefits of using models to produce rapid decision-making guidance for COVID-19 control in the WHO Eastern Mediterranean Region (EMR), as well as challenges that we have experienced regarding conveying uncertainty associated with model results, synthesizing and comparing results across multiple modelling approaches, and modelling fragile and conflict-affected states.

## Introduction

In January 2020, a cluster of patients with pneumonia of unknown cause was reported in Wuhan, China; the disease, later named COVID-19, was found to be due to a novel coronavirus, the severe acute respiratory syndrome coronavirus 2 (SARS-CoV-2). The disease spread worldwide and was declared a pandemic by the World Health Organization (WHO) on 11 March 2020 (1). In the EMR, the first cases were detected in the United Arab Emirates (UAE) on 29 January 2020. By 15 January 2021,, the 22 countries and territories of the Region have been affected with more than 4.5 million reported cases and almost 130 thousand reported deaths (2). However, the true burden of COVID-19 in the Region is not fully measured due to various factors including limiting testing and inadequate surveillance systems in many countries. The number of cases of COVID-19, and attributable deaths, is under-reported to an unknown extent. Several EMR countries suffer from economic insecurity, armed conflict and political instability, and have limited capacity for public health surveillance (3).

Since the beginning of the pandemic, public health authorities have faced an urgent need to make decisions regarding various actions to mitigate the spread of the virus and the associated pressure on healthcare systems. Early detection and isolation of cases, contact tracing and quarantine of contacts are critical to prevent further spread of the virus and reduce morbidity and mortality (4,5). Also, early in the outbreak, many countries introduced additional public health and social measures (PHSM), which include intervention strategies such as movement restrictions, closure of schools and businesses, and international travel restrictions to slow down disease spread and relieve some pressure on clinical care services (6). By mid-March 2020, all countries in the EMR had applied some form of PHSM. Not only are these measures difficult to implement in some countries of the region, but they are also accompanied by considerable social and economic costs. Thus, decisions to tighten, loosen, or re-instate PHSM should be based on local epidemiological data, risk assessments (7) and available scientific evidence (8).

### Applying mathematical models to support decision-making in the Covid-19 pandemic

Mathematical epidemiological models are useful for informing such decisions through exploring and understanding the consequences of a variety of plausible scenarios regarding assumptions about virus and disease characteristics, health system capacity, and the timing and strength of PHSM (9–11). We provide a background to how epidemiological models have been applied to analysing the COVID-19 pandemic in the Appendices (Text A1). In summary, mathematical models allow prediction and comparison of key epidemiological outcomes such as the daily incidence of infections (including severe infections that require hospitalisation and intensive care) and deaths. There is considerable uncertainty associated with these predictions, particularly as COVID-19 is a novel viral disease in humans, for which the evidence on transmission and clinical outcomes continues to evolve, and as such many of the parameters and processes that govern its spread and clinical outcomes are unknown or poorly quantified (10–12). Moreover, models make simplifying assumptions in their representation of reality, and they evolve and adapt over time with the emergence of new data and evidence (12). Nonetheless, the qualitative behaviour of models can effectively address certain questions of current importance to policy-makers. For example, models can compare different scenarios that represent alternative PHSM strategies and give insight into their relative benefits (10,13), such as flattening the epidemic curve and reducing the expected burden on the healthcare system (14). Models can warn about future waves of infection, subject to their assumptions about infection, re-infection and immunity (15). Importantly, modellers need to work closely with decision-makers and the media to ensure that model predictions are interpreted and used appropriately, that the associated uncertainty is clearly communicated (12), and that the context and motivation of the developers and users is transparent (10). This collaboration is also important for collection of model inputs as the reliability of any prediction is conditional to the quality of the epidemiological data feeding the model. Moreover, while a model is a useful tool for supporting decision-making, its predictions should be interpreted across a realm of other tools and data sources.

### Participatory approach to modelling the Covid-19 pandemic in the EMR

Public health officials in EMR countries noted how mathematical epidemiological models were informing public health decisions in the countries that were hit earlier in the pandemic (16,17). As a result, some of them have expressed an interest in receiving technical support from WHO for mathematical modelling to interpret the potential epidemiological spread of the outbreak, given different PHSM scenarios. Accordingly, the WHO Eastern Mediterranean Regional office (EMRO) established a modelling support team in mid-March as part of the Information Management pillar in the COVID-19 Incident Management Support Team (IMST) (18). This is the first time the WHO Regional Office utilized modelling as a tool in an ongoing outbreak response. Here we present an innovative approach that was developed by the EMRO modelling support team to conduct participatory modelling analyses together with public health professionals from the EMR in order to support decision-making.

The primary objective of the modelling support team was to address the imminent decision-making needs faced by policy-makers. Another objective was to provide training on mathematical epidemiological modelling, including the underlying concepts and methods, in order to promote awareness of how models work, how they can be used effectively, and to work towards building technical capacity for mathematical modelling in the EMR.

A participatory approach was adopted in order to conduct country-specific modelling analyses that were relevant to the PHSM being considered for the country. As part of this approach, policy-makers from the Ministries of Health in the countries were actively involved throughout all stages of the modelling process (13,19,20). The country team also often included WHO Country Offices, WHO Headquarters, researchers from academia and other UN agencies (see Figure A3 in the Appendices). Participatory modelling can facilitate the translation of model findings into decision-making in a number of ways. Firstly, it can guide the modelling analysis to identify and address questions that are relevant and important to public health in the country. It can also encourage shared learning amongst researchers, modellers and policy makers, and foster common understanding and expectations (19). Importantly, the involvement of within-country analysts and policy makers builds trust, ownership and support (19,20), meaning that the findings of the analyses will be more likely to be considered in decision-making.

### Policy questions posed by policy-makers in EMR

After consulting with policy makers from countries, by the end of March 2020, the EMRO modelling support team identified the most urgent questions to address through modelling exercises: (i) how many infections and deaths are expected at different times; (ii) what are the expected healthcare system requirements and (iii) what is the effect of PHSM on the spread of the epidemic? In order to be suitable for supporting rapid decision-making in countries in the EMR, many of which are Low and Middle Income Countries (LMICs), the modelling tools used needed to be readily adaptable to country-specific settings and PHSM policies, with data inputs swiftly attainable and requiring low computational resources. It was also necessary to use an interpretable modelling approach that could be readily communicated in non-technical terms to participants from a variety of disciplines.

### Application of the CoMo Model in the EMR

The team found that the CoMo model developed by the COVID-19 International Modelling Consortium (CoMo Consortium) (21) offered a user-friendly interface to modelling impacts of PHSM on the time trajectories of infections (including severe infections requiring hospitalisation and intensive care) and deaths. The CoMo model was able to address the questions posed by policy makers in the region, had the flexibility to be adapted to the regional context, and had a computationally efficient online application which allowed the model to run quickly on large populations.

The CoMo model (21) is an age-dependent deterministic SEIR (Susceptible -Exposed -Infectious – Recovered) model that predicts daily incidence of COVID-19 infections in the population under different assumptions about PHSM. Details of the CoMo modelling methodology are provided in the Appendices (Text A1). In summary, the model considers five levels of infection severity: asymptomatic infections, symptomatic infections, and infections requiring hospitalisation, intensive care treatment and ventilated intensive care treatment. Infection severity and associated mortality are age-dependent, in that the proportion of infected individuals requiring hospitalisation, and the proportion that die, varies with age. The CoMo model incorporates a hospital sub-model that indicates when hospital treatment requirements, including treatment in hospital beds, ICUs and ventilators exceed the capacity of the country’s healthcare system.

The CoMo model incorporates an explicit representation of the PHSM that have been commonly used to mitigate the spread of COVID-19. The model considers PHSM that target infected individuals, including self-isolation of symptomatic individuals and self-quarantine of members of their household, screening of the contacts of individuals with a positive diagnostic test result, and mass testing. The model also considers PHSM that aim to limit transmission throughout the wider population, including school closure, workplace closure, physical distancing measures, border closure, shielding elderly individuals, hand-washing and mask-wearing. For each intervention, the user provides inputs about the coverage, defined as the proportion of the total population to which the intervention applies. The user also inputs the intervention timing and duration, and the adherence of the population to the intervention. These coverage and adherence parameters act on age-specific social contact rates in home, work, school and other community environments, which are estimated in the model using the country-specific contact matrices developed by Prem et al. (23). A useful feature of the CoMo model is that it allows the coverage of each intervention to vary over time, meaning that the intervention can have a stronger effect over some time periods and a weaker effect over other time periods. We have used this feature in applying the model to EMR countries to represent relaxations of PHSM (such as during Eid and Ramadan periods) and strengthening of measures in response to resurgences in infections.

### Participatory Modelling Process

The EMRO modelling support team, together with the CoMo Consortium, has been adapting the CoMo model to investigate COVID-19 spread dynamics in countries in the EMR, as well as interpreting the results and conveying the models’ caveats to the countries. Throughout this process, the team has assisted countries in identifying data sources, gathering country-specific data, documenting model outputs and preparing policy briefs. The team supports capacity building by delivering training and demonstrations of the models and the online tool. Establishing participatory collaborations with EMR countries to adapt and run the CoMo model involved the following stepwise approach:

1. *Introduce modelling approach:* The EMRO modelling support team introduces the modelling approach to the interested countries using an audio-visual presentation that provides a non-technical overview of the CoMo model, including the main inputs, outputs and limitations. This is supported by social media videos in three regional languages (24–26). The introductory sessions and the videos convey basic information about epidemiological models and the uncertainty about model parameters and structure.
2. *Assemble collaborating team:* A team of core collaborators that will participate in the modelling analysis is assembled. This includes public health professionals from the country Ministry of Health and the WHO Country Office as well as the EMRO modelling support team.
3. *Collect model input data:* Once the country embarks on a collaboration, the data collection template and a reference manual describing the model parameters and their definitions are shared. The country is supported during the collection of country-specific model parameters by convening data collection webinars and sharing data sources.
4. *Review input data:* The EMRO modelling support team corroborates the data inputs by cross-checking with published literature and the country. The team also cross-checks the assumptions being made about the strength and timing of the PHSM implemented in the country with the Google mobility reports (27) and Oxford government response tracker (28). Any discrepancies are discussed with collaborators from the country to develop a common understanding and agree on any revisions.
5. *Conduct initial modelling analysis:* Modelers from the EMRO modelling support team and the country conduct an initial modelling analysis to produce preliminary results. The analysis employs a scenario-based approach that develops customised scenarios which compare different PHSM implementation strategies based on the questions posed by the countries. The team drafts a technical report containing details about model inputs, methods, results and limitations that is then shared with Member State collaborating team.
6. *Review initial results:*The country collaborating team reviews the technical report and discusses with the EMRO modelling support team any modifications or updates required.
7. *Update modelling analysis:* The EMRO modelling support team updates the modelling analysis and produces an updated technical report which is shared with the country collaborating team.
8. *Summarise policy implications:*The EMRO modelling support team prepares a non-technical audio-visual presentation together with a policy brief document that target a wide public health audience and interpret the main findings of the analysis from the perspective of decision-making, as well as conveying the uncertainty and limitations.
9. *Continue collaboration:* Engagement with countries continues as required, to update the modelling analysis according to the progression of the COVID-19 outbreak and the control strategies under consideration in the country.

The components of this participatory modelling approach, including participants, resource requirements, and methodological approaches, are summarised in Table A1 and Figure A3 of the Appendices. We have evaluated our experience of implementing the above nine step process to identify benefits and limitations, and provide recommendations for the continued use of this approach (Table A2 of the Appendices).

### Progress to date

As of mid-January 2021, eleven countries, namely Afghanistan, Djibouti, Egypt, Iraq, Jordan, Lebanon, occupied Palestinian territories (oPt), Pakistan, Syria, Tunisia and Yemen have been engaged in collaborative modelling with EMRO. Of these, seven, namely Afghanistan, Jordan, Lebanon, Pakistan, Egypt, oPt, and Tunisia had reached stage eight of the above process and disseminated the results of the modelling analyses and policy implications to the decision-makers. Scientific committees and decision makers in some of those countries are consulting the model’ s projections, in conjunction with the epidemiological data and other data sources, when examining current policies in place and weighing possible options for instating or loosing certain measures. For instance, Tunisia’ s epidemiologists are exploring options of immediate or gradual release of physical distancing and working from home measures (29). The EMRO modelling support team is currently in the process of documenting examples where model results have already been incorporated into policy. One example is the school opening strategies, such as the cyclical school opening strategy adopted in oPt (30). Another example is the implementation of one day lockdown per week and a daily night time curfew in addition to keeping schools online till the end of the semester in Jordan (31). Investigating testing strategies and possibilities of underreporting with the modelling tool is also currently being explored in some countries.

### Challenges

There are a number of barriers to effective and widespread use of models in the EMR such as the insufficient availability of national data and limited national modelling capacity. There is considerable uncertainty associated with modelling the dynamics of novel viruses (10,11), and interpreting the results can be challenging given the general limitations of the models and the input data on which they are based (Text A1 and Table A2 of the Appendices). The EMRO modelling support team has tackled these issues by communicating throughout all stages of the modelling collaboration the expectations about the appropriate uses and limitations of models. The social media videos that were developed by the team are presented in Arabic, English and French, and describe the uncertainty associated with epidemiological models of COVID-19 and how model results can be interpreted to guide decision making (24–26). These videos were disseminated on different platforms and at the time of writing had received 25359 views across all platforms. The team also encourages national capacity building by providing training on the modelling approach and caveats, as well as on using the online software and the interpretation of results. These challenges can be further addressed through encouraging active engagement in modelling as a part of public health decision-making as well as strengthening the local systems for epidemiological surveillance and data storage.

At present, several modelling analyses of the COVID-19 pandemic have been conducted for EMR countries by different international research groups, each using different modelling approaches and making different assumptions about viral transmissibility and severity, and the implementation of PHSM (32–36). This can lead to discordant sets of results and guidance being presented to policy makers which can be difficult to assimilate, confusing, and potentially leading to a distrust, and loss of interest, in models. A challenge for the modelling community lies in systematically comparing and synthesizing results provided by different models in order to provide clear and consistent guidance to decision-makers (37). Model comparison initiatives that are currently reviewing the results of multiple COVID-19 epidemiological models are a promising step towards achieving this goal (38,39). The EMRO modelling support team is involved in these activities through an international network of collaborations with academic institutions and other UN agencies (Figure A3 of the Appendices).

The socioeconomic and geopolitical situation in the EMR is another unique challenge, not only for implementing PHSM and but also for modelling the COVID-19 outbreak. More than half its countries are currently suffering from armed conflict and political instability (40). Additionally, it is estimated that out of the 600 million people living in the region, 13 million are internally displaced, 12 million are refugees (41), and 46 million are low income labour migrants (42). These populations often reside in overcrowded spaces with poor living conditions, such as camps or camp-like settings, where it can be difficult to apply certain public health measures, such as physical distancing and self isolation. Basic personal protective measures (such as hand washing) adherence is problematic as well due a lack of basic sanitation services. However, despite this, no ‘explosive outbreaks’ were reported from countries affected by conflict and humanitarian emergencies and the attack rates appear to be substantially lower than other countries in the region (3). A number of approaches for modelling COVID-19 dynamics in IDPs and refugee camps are being developed (e.g. (43)) and the EMRO modelling support team is investigating the application of such *approaches to the region*.

### Conclusion and way forward

Mathematical epidemiological models offer a whole system approach to analysing the COVID-19 pandemic and can assist countries with the multifaceted aspects of public health planning and decision-making. The EMRO modelling support team is a new initiative for the WHO, and it has developed a participatory modelling approach for the EMR in order to undertake analyses that are tailored to the context of the countries in the region. The strength of the approach lies firstly in the rapid and active engagement of public health policy makers with epidemiologists and modellers from countries within the region. This was achieved through coordinating with other WHO regional offices, EMR WHO country offices, and WHO headquarters. Secondly, the team’ s activities continue to benefit from establishing collaborations with academic and other UN institutions, which has provided access to technical expertise covering a wide range of modelling approaches. Through this participatory approach, the model will continue to be revised based on prevailing decision-making needs, such as the recent added function of investigating various scenarios of COVID-19 vaccine introduction. The quality of model results, however, depends on the quality of the epidemiological data that serve as model inputs, and model projections are not a replacement for a lack of comprehensive surveillance and experimental evidence. The challenges associated with mathematical modelling are pronounced in the EMR, where several member states do not have sufficient resources and technical capacity for collecting, processing and modelling epidemiological data. Data unavailability and poor quality, especially in conflict-affected countries, will increase the uncertainty associated with model findings. The participatory modelling approach, webinars, social media videos, technical reports and policy briefs are steps towards building capacity for mathematical modelling in EMR, raising the awareness of the use of mathematical models, their benefits and limitations. By continuing active engagement with country collaborators, and the international modelling community, we will continue to strengthen epidemiological surveillance and modelling analysis for public health decision-making in the region.

## Data Availability

There is no data repository associated with this manuscript

