## Supplementary material for "A participatory modelling approach for investigating the spread of COVID-19 in countries of the Eastern Mediterranean Region to support public health decision-making"

Text A1: Applications of Susceptible-Exposed-Infectious-Recovered (SEIR) models to guide policy making throughout the COVID-19 pandemic…………………..2

Table A1: Summary of the methodological approach, including participants and resource requirements, for the participatory modelling analyses conducted by the WHO EMRO modelling support team……………………………………………………..…..7

Table A2: Evaluation of the modelling process and results……………………………..8

Figure A3: Participatory modelling approach structure and participants………...9

### Text A1: Applications of Susceptible-Exposed-Infectious-Recovered (SEIR) models to guide policy making throughout the COVID-19 pandemic

Mathematical epidemiological models have been widely applied throughout the COVID-19 pandemic to provide quantitative and reproducible in silico simulation of the epidemiological dynamics of COVID-19. They provide a holistic approach to modelling the epidemiological system, considering multiple dynamic processes that interact to drive virus transmission patterns. For example, they model processes of viral transmission in the human population, utilising data describing expected rates of human-to-human contact in different environments (such as workplace, school and home environments) and different geographic regions. They also include data on the characteristics of the COVID-19 virus such as its transmissibility and the duration of the viral incubation period, as well as the expected rates of severe infections and deaths. They can include information about how these processes vary across different demographic variables and vulnerability characteristics. Models combine these epidemiological processes and describe their highly non-linear interactions, which are difficult to understand using only intuition (1). Models have been applied globally throughout the pandemic to guide policy makers about possible future trajectories of COVID-19 infections and deaths, the consequent demands on healthcare systems, and how these can be influenced by the implementation of various public health and social measures (PHSM).

#### 1.1 The SEIR model

Many epidemiological models of COVID-19, including the CoMo model, are based on mechanistic Susceptible-Exposed-Infectious-Recovered (SEIR) modelling approaches (2) that model the mechanisms of viral transmission and the progression of the infection in individuals who have contracted the virus. In its simplest form, the SEIR model subdivides the human population into four classes of individuals (Figure A1): (i) Susceptible individuals, meaning those who are not currently infected and can potentially contract the infection upon making an infectious contact; (ii) Exposed individuals, meaning those who have become infected with a virus that is still undergoing incubation; (iii) Infectious individuals, meaning those who are infected with a virus that has completed the incubation phase and can potentially cause clinical symptoms; (iv) Recovered individuals, meaning those who have recovered from the infection and are no longer infected. The rates of transition between each of the four classes are defined by parameters that are estimated based on observations of viral spread and infection progression in the human population. Importantly, the rate at which Susceptible individuals become infected is proportional to the number of individuals who are Infectious (Figure A1), thus infections grow exponentially in the initial phase of the epidemic. The mathematical model tracks the temporal evolution of the number of individuals in each of the Susceptible, Exposed, Infectious and Recovered classes using a series of differential equations.
The SEIR framework can be modified to include several types of additional structure that is relevant to the COVID-19 epidemic. For example, the CoMo model subdivides the population according to age (Figure A2), and allows patterns of human-to-human contact to vary with age as well as environmental setting. Age-dependent contact rates are specified for four different

environments, including home, school, workplace and other environments, where the "other" environment type summarizes all contacts outside the home that are not workplace or school contacts. The CoMo model allows the impacts of specific PHSM on different contact rates to be explored, including those that affect specific environments such as the closure of schools or workplaces, as well as wider lockdowns that reduce contacts in all non-home environments. Additionally, the model can consider reductions in contacts in individuals who are self-isolating or who are in quarantine. It is important to note that human mobility, and the spatial and geographic dimensions of human contact patterns, are also an important determinant of the spread of COVID-19 infections. At present, the CoMo model does not include spatial structure, and therefore does not represent variation in contact rates and viral transmission according to the spatial proximity of individuals, or across different geographic regions. There are other SEIR models of the COVID-19 pandemic that have been applied to Lower and Middle Income Countries (LMICs) that do explicitly model spatial structure and human movement (3).

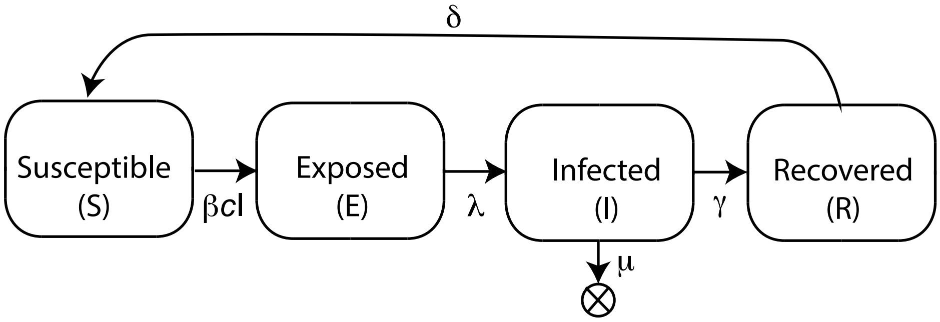


**Figure A1.** A simple SEIR model. Susceptible individuals contract the infection and become Exposed at a rate proportional to the number of infected individuals in the population, *I*, the average number of daily contacts, *c*, and the probability of viral transmission given an infectious contact, *β*. Exposed individuals enter the Infected class at a rate λ, and infected individuals either recover at a rate γ or die from the disease at a rate μ. Recovered individuals lose their immunity to reinfection at a rate δ.

The CoMo model also incorporates a hospital sub-model (Figure A2) that subdivides infectious individuals according to hospital treatment requirements, and represents an age-dependent increase in the likelihood of severe infections that require specialized hospital treatment. This allows projection of the expected burden on the healthcare system. As we learn more about the COVID-19 virus, models can be further refined to represent additional mechanisms and structure. For example, recent updates to the CoMo model consider varying degrees of immunity to re-infection with COVID-19 in recovered individuals and how this depends on the decay of COVID-19 antibodies over time (4). Models may also consider how the disease burden is impacted by the prevalence of other health conditions that can increase vulnerability to severe disease outcomes.

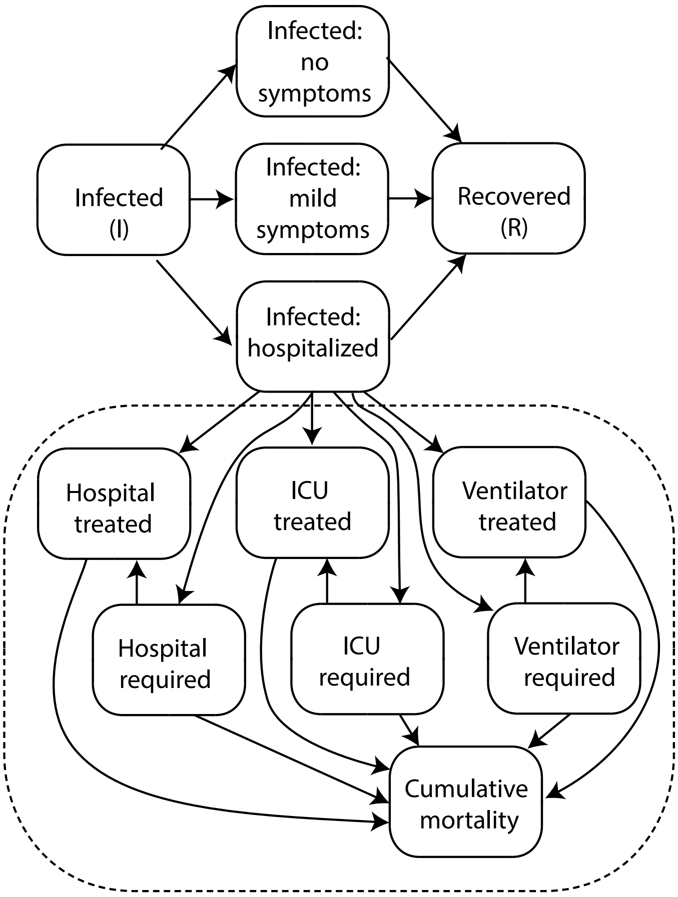


**Figure A2.** The CoMo model incorporates a hospital submodel (enclosed by the dashed line). The model subdivides the Infected individuals into those who experience no symptoms, mild symptoms, or a level of severity that requires hospitalized treatment. Hospitalized individuals are categorized according to the type of treatment they require (hospital, ICU, or ICU and Ventilator treatment) and whether or not they are receiving the required treatment. When the hospital reaches its capacity for a given treatment type, individuals who require a given treatment but do not receive treatment are placed in either the "Hospital required", "ICU required", or "Ventilator required" classes according to the type of treatment required. These individuals can move into the "Hospital treated", "ICU treated" or "Ventilator treated" classes if resources become available. COVID-19 attributable mortality occurs only in individuals with an infection severity that requires either hospital, ICU or ventilator treatment.

**1.2. Appropriate use of models to guide policy-making**

The COVID-19 pandemic has generated a heightened interest in mathematical epidemiological models across many sectors of society as policy-makers and the public seek clarity around the future implications of this novel virus. Mathematical modelling has played a prominent role in policy making throughout the pandemic, and this novel focus on modelling, together with the novelty of the pandemic itself, has led at times to confusion and misunderstanding about how to use models appropriately and effectively to guide public health decision making. The scientific modelling community has an important role to play in guiding public health managers, politicians and the wider public about how to interpret and apply the results of models. Since the beginning of the pandemic several modelling research groups have published guidance that summarizes and reviews, in non-technical language, key underlying principles for using mathematical models to guide policy making (1,5,6).

An important and widely emphasized principle is that model predictions need to be accompanied by a full and transparent assessment of the associated *uncertainty* (1,5–8). Predictions need to be presented as range of values describing the uncertainty interval, namely the confidence or credible interval (CI), rather than a single numeric value. CIs only provide estimates of a part of the uncertainty, however, and do not capture all uncertain aspects, because all models are a simplified representation of reality and are based on assumptions. It is therefore important that statements about uncertainty include a full articulation and assessment of model assumptions, including how these may impact model predictions, and that areas of ignorance are acknowledged.

Moreover, the validity of model estimates depends entirely on the quality of the epidemiological data used to construct and parameterize the model. In the case of COVID-19, there is considerable uncertainty in reported epidemic trajectories due to a multitude of factors such as biases in the sets of individuals who are tested, under-reporting of cases and deaths, and uncertainty in laboratory assays and diagnoses. Key aspects of the epidemiology of COVID-19 remain uncertain, including the extent and duration of immunity following infection, the extent and transmissibility of asymptomatic infections, and patterns of human-to-human contacts across different populations and regions (1).

Thus, while public health managers seek accurate numbers about future COVID-19 infection rates, hospitalizations and deaths, models cannot provide predictions with this level of certainty. Models are not crystal balls (9), and it is not appropriate to use models to provide a single precise forecast of future epidemiological outcomes (6). Models are more effective when used to assess relative impacts across several sets of predictions (6). For example *scenario-based* modelling approaches present results across multiple scenarios that make different explicit assumptions about parameters and processes of interest (10,11), and draw insights from the qualitative as well as quantitative differences in predictions across different scenarios. From the perspective of public health policy making, it is useful to focus on the relative impacts of different PHSM, and to include an assessment of the scale of the expected burden on the healthcare system (6).

Quantitative predictions are typically only useful in short term forecasts. An important aspect of assessing model validity and uncertainty involves follow-up analyses to compare short term predictions to observations as they become available. It is critical that modelling analyses are *reproducible* so that predictions made at different times can be regenerated and assessed.

There can be a tendency to favour complex models over simpler approaches, and to assume that greater complexity leads to more realistic and robust predictions. This will not be the case, however, if complex models misrepresent or omit key biological aspects (1,6). It is therefore important that model choice and design incorporates an assessment of the *trade-off between simplicity and complexity*. This trade-off depends on the availability of data to support the development of a more complex model. Modellers need to acknowledge issues associated with parameter identifiability, and recognize that it is more difficult to infer parameters, and to identify errors, for complex models than for simpler models.

Moreover, it is beneficial to use modelling approaches that are *interpretable* to both analysts and end users, as this allows an assessment of how model results are related to the choice of questions addressed, the inputs used, and the assumptions made (5). This feedback is valuable to adapting and improving subsequent modelling analyses, and continuing to guide decision-making.

Finally, it is important that modelling analyses are accompanied by a *description of the context* in which the analyses were conducted, including the intended purpose of the analyses, and the background and motivation of the model developers, analysts and stakeholders. The choice of modelling approach and the design of analyses is never neutral (5), and can greatly influence the results and conclusions. Analysts therefore need to provide an explicit statement of these biases and motivations. Scientists need to work with politicians, journalists and the media to ensure that model results are not politicized, and reports of model predictions are couched in appropriate statements about context, the key caveats and uncertainty.

**Table A1: Summary of the methodological approach, including participants and resource requirements, for the participatory modelling analyses conducted by the WHO EMRO modelling support team.**

| Participants | Resources required | Methods | |
|---|---|---|---|
| | | Considerations in choosing the modelling approach | Guiding principles for modelling analysis |
| • Ministry of Health in Member States<br>• WHO focal points<br>• Technical modellers<br>• Surveillance officers<br>• Epidemiologists<br>• Communication and Policy experts | • Software application for implementing epidemiological modelling analysis that is:<br>(i) computationally efficient<br>(ii) user-friendly<br>(iii) reproducible<br>• Additional mathematical and statistical software for analysis and visualization of model results e.g. R, python.<br>• Software for producing scientific presentations, reports and publications e.g. MS Office, Reference Manager<br>• Software for telecommunication support e.g. Cisco Webex<br>• Access to WHO and publicly available databases, including WHO dashboards, GIS databases, Google mobility databases and academic research databases. | • Timely availability for immediate application<br>• Open source, user-friendly software application<br>• Access to technical support from model developers<br>• Model and software is actively developed and maintained<br>• Model is adaptable to LMIC settings<br>• Access to an active, collaborative consortium of model users and developers<br>• Modelling approach is transparent and interpretable to users from a wide range of professional backgrounds. | We follow published scientific advice summarized in **Text A1** for applying mathematical models to analyzing the epidemiological dynamics of COVID-19. In summary this advice recommends:<br>• full and transparent assessment of model uncertainty<br>• present results for several equally plausible scenarios<br>• all results must be reproducible<br>• assessment of tradeoffs between simplicity and complexity<br>• use an interpretable modelling approach and discuss results in terms of assumptions and limitations.<br>• Provide a description of the context and motivations for conducting the modelling analyses |

**Figure A3: Participatory modelling approach structure and participants**

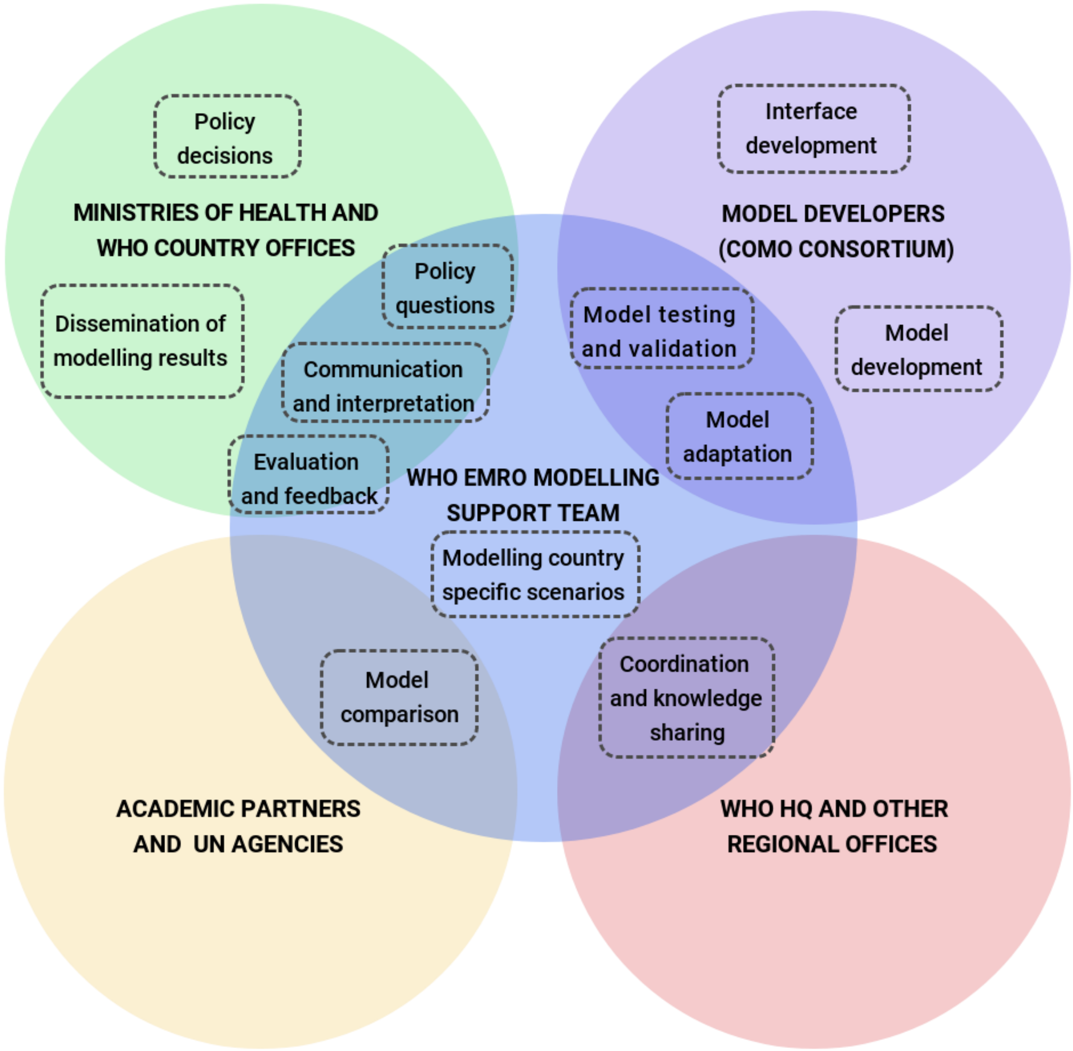

**Table A2: Evaluation of the modelling process and results**

| Process and results | Evaluation questions | Summary of experience |
|---|---|---|
| **1. Introduce modelling approach** | - Do the participants understand and agree on expectations regarding what the modelling analysis can achieve? | - At the start of the process, it is important to establish a common expectations and understanding about what models can and can not do. |
| **2. Assemble modelling team** | - Does the modelling team has a representation from an appropriate range of backgrounds? | - The modelling team should consider participants from broad range of backgrounds as detailed in (table A1) and (figure A3) |
| **3. Collect model input data**<br><br>**4. Review input data** | - Could the necessary country-specific model inputs be collected and validated? | - Unavailability and poor quality data especially from conflict affected countries<br>- Hospital line lists are frequently not sufficient to fully quantify the rates of hospitalisation and deaths in the population<br>- Not sufficient community level data to fully quantify the effect of public health and social measure on the virus transmission rate<br>- These sources of uncertainty should be recognised<br>- A scenario-based modelling approach (text A1) needs to be used to provide model results across range of possible parameter values<br><br>- After model inputs are being cross-checked against literature values and other data sources, the modelling team should discuss any discrepancies and have common understanding and a consensus on the input values prior to conducting any modelling analysis. |
| **5. Conduct initial modelling analysis** | - Do country collaborators have the capacity to run models independently? | - Several EMR countries has limited capacity for data processing and modelling<br>- The participatory modelling process is a step towards building national modelling capacity |

| Process and results | Evaluation questions | Summary of experience |
|---|---|---|
| **6. Review initial results** | - Can modelling results be interpreted in the light of uncertainties around model inputs and model limitations? | - Model results interpretation can be challenging given the limitations of the models and the quality of the input data<br>- Continuous communication is needed to clearly interpret model results and convey models limitations and caveats |
| **7. Update modelling analysis** | - How does the modelling team plan to update the modelling analysis? | - Models need to be regularly updated (e.g. monthly) to produce short-term forecasts<br>- It is important to conduct regular literature searches to guide updating model inputs and/or structure<br>- Updating model parameters should be after developing a common understanding of model's main findings and be subjected to any changes in disease dynamics and/or the implementation of PHSM in real life.<br>- Timely to respond to pressing policy questions (e.g. different school re-opening strategies) |
| **8. Summarise policy implications** | - Are the policy implications clearly stated and can they be translated into actionable decisions? | - Policy implications should be clearly and concisely summarised in plain language and target a wide range of backgrounds<br>- The trust and ownership established through the participatory approach facilitates the translation of model results and their policy implications to decisions |
| **9. Continue collaboration** | - What are the key drivers to continue collaboration? | - Continuing collaboration builds capacity for epidemiological modelling in the region<br>- The participatory approach encourages continued involvement and collaboration |
